# OpenScientist: evaluating an open agentic AI co-scientist to accelerate biomedical discovery

**DOI:** 10.64898/2026.03.15.26348338

**Authors:** Kaleigh F. Roberts, Zachary B. Abrams, Luca Cappelletti, Mahdi Moqri, Nicholas Heugel, J. Harry Caufield, Mathieu Bourdenx, Yan Li, Jineta Banerjee, Luca Foschini, Diego Galeano, Nomi L. Harris, Melody Li, Kejun Ying, Justin A. Melendez, Nicolas R. Barthélemy, James G. Bollinger, Yingxin He, Vitaliy Ovod, Tammie L. S. Benzinger, Shaney Flores, Brian A. Gordon, Adegoke A. Ojewole, Mukta Phatak, Donald L. Elbert, Sarah Biber, Eric C. Landsness, Christopher J. Mungall, Randall J. Bateman, Justin T. Reese

## Abstract

**Background:** Advances in medicine depend on analyzing large and complex data sources, but discovery is partly constrained by the limited time and domain expertise of human researchers. Agentic artificial intelligence (agentic AI) can accelerate discovery by automating components of the scientific workflow, including information retrieval, data analysis, and knowledge synthesis.

**Aim:** OpenScientist, an open-source agentic AI co-scientist, aims to accelerate biomedical discovery by semi-autonomously investigating scientist-defined queries and generating clinically relevant, verifiable scientific insights.

**Methods:** Domain experts evaluated OpenScientist for novel discoveries in four clinical case studies: (1) a prespecified analysis in a community-based Alzheimer’s disease biomarker cohort, (2) unsupervised modeling for plasma proteomic survival prediction, (3) hypothesis investigation in single-cell transcriptomic data from neurons with neurofibrillary tangles, and (4) hypothesis generation with validation in a multiple myeloma dataset with a randomized negative control.

**Results:** OpenScientist completed analyses in minutes that otherwise would take weeks to months of human time and expertise. It identified %ptau217 as the best predictor of amyloid PET status, generated a plasma proteomic survival model with performance comparable to published models, proposed a mechanism linking tau pathology to altered lysosomal acidification, and generated multiple myeloma hypotheses that were validated in an external cohort while distinguishing true signal from randomized controls.

**Conclusion:** OpenScientist demonstrates that open, auditable, agentic AI can support real-world clinical research by generating hypotheses, executing analyses, and discovering insights from complex datasets.

## Introduction

The accelerating pace of biomedical data generation has created an urgent need and opportunity for computational tools that can autonomously extract mechanistic insights from complex datasets. Recent advances in large language models (LLMs) and in particular agentic methods have enabled the development of AI-powered research assistants capable of generating hypotheses, designing experiments, and interpreting results. However, most existing AI scientist platforms remain proprietary, limiting reproducibility and broader adoption within the scientific community.

AI models have demonstrated impressive capabilities across scientific domains, exemplified by Google DeepMind’s GraphCast for weather prediction and AlphaFold for protein structure determination.^1,2^ More recently, a new class of AI scientific assistants explicitly targeting research workflows has emerged, including Google’s Co-Scientist^3^, Sakana’s AI Scientist^4^, Edison Scientific’s Kosmos^5^, Biomni^6^, K-Dense^7^, and many others, each designed to support aspects of hypothesis generation, multimodal data interpretation, and scientific reasoning.^8–10^ In parallel, agentic coding systems such as Anthropic’s Claude Code and OpenAI’s Codex have demonstrated the ability to write, debug, and execute scientific analysis code autonomously. Despite these advances, a critical gap remains: existing platforms are predominantly closed-source, preventing independent verification, customization for domain-specific workflows, and integration with institutional computational infrastructure.

Here, we present OpenScientist, an open-source autonomous discovery platform designed to democratize AI-assisted scientific research (**Figure 1**). Unlike proprietary systems, OpenScientist provides complete transparency in its hypothesis generation workflow, enabling researchers to understand, validate, and modify

**Figure 1.**
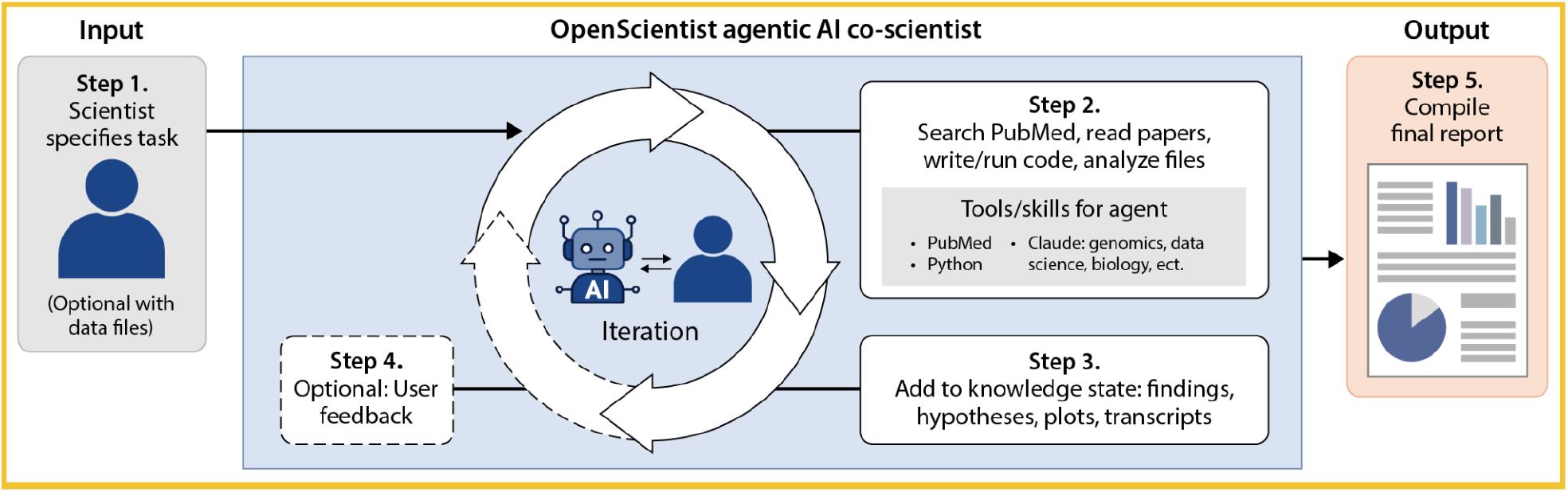
Overview of the OpenScientist scientific investigation workflow. 1) Scientist describes their scientific task (a query, a scientific analysis, etc.), and optionally includes one or more data files to support the task. 2) The agent is given the task and data files, and autonomously investigates by querying PubMed, examining papers, writing and executing code, guided by a suite of Agent Skills, comprising advice in various scientific domains for the agent. 3) The artifacts from the investigation, including finding, hypotheses, analyses, plots, are saved in a knowledge state data structure (KSDS). 4) Optionally (if the user selected co-investigator mode), feedback from the user is collected and stored in the KSDS. The new iteration of OpenScientist then begins (step 2), with the KSDS provided to build on existing knowledge. 5) After N iterations (10 by default), the results are compiled into a final report.

the AI’s reasoning process. The platform is largely agnostic to data format and data types, and accepts diverse data types such as electronic health records, genomics, transcriptomics, proteomics, metabolomics, structural biology, histopathology, imaging, and biomarker data, in multiple input formats including text, tabular data, binary omics formats, and images. OpenScientist autonomously formulates testable hypotheses, executes computational analyses, extracts biological and biomedical findings, and produces mechanistic insights grounded in data and published literature.

OpenScientist’s architecture leverages Agent Skills and the Model Context Protocol (MCP) to provide domain-specific tools for data analysis, literature search, and knowledge synthesis. The system operates through an iterative discovery loop: in each iteration, OpenScientist generates hypotheses based on accumulated knowledge, designs and executes computational experiments using sandboxed Python code execution, queries PubMed for context, and updates its knowledge state with validated findings. This approach mirrors the scientific method while maintaining full auditability through state tracking and transparent logging of all analyses.

A key feature of OpenScientist is its modular skills system, which separates domain-agnostic workflow skills (hypothesis generation, result interpretation, prioritization, stopping criteria) from domain-specific analytical skills (metabolomics, genomics/transcriptomics, structural biology, statistics). This architecture allows researchers to extend OpenScientist’s capabilities for specialized applications without modifying core system logic. The platform supports multiple LLM providers including Anthropic, Google Vertex AI, AWS Bedrock, Azure Foundry, and CBORG (an internal compute resource at Lawrence Berkeley National Laboratory).

We demonstrate OpenScientist’s capabilities across multiple clinical and biomedical domains and discuss how its open-source nature addresses critical needs for transparency, reproducibility, and customization in AI-assisted scientific discovery. By providing a fully auditable, extensible platform for autonomous hypothesis testing, OpenScientist aims to accelerate discovery while maintaining the scientific rigor essential for biomedical research.

## Methods

### System Design and Architecture

OpenScientist operates as an autonomous discovery loop that iteratively refines hypotheses through computational analysis and literature integration (**Figure 1**). OpenScientist is built using Claude Code with a design that allows future support for alternative AI agents, and a public collection of Agent Skills (https://github.com/K-Dense-AI/claude-scientific-skills) for specific biomedical knowledge domains, analytical functions, or bioinformatics tools.^11^ Claude Sonnet 4.5 was used in all OpenScientist runs in this manuscript. These skills encapsulate capabilities such as statistical analysis, literature retrieval, knowledge graph reasoning, and biomedical interpretation. The system runs within a Docker container for portability and reproducibility, and is orchestrated by an agentic controller that dynamically selects and combines skills based on the user’s query and the evolving state of the investigation.

Scientists initiate an analysis by entering a free-text query describing the scientific task, optionally including one or more datasets to guide the investigation, as well as any supplementary reference material (articles, slide decks, etc). Once initialized, OpenScientist autonomously executes an iterative analysis loop. The system performs N iterations of research cycles, each comprising a set of autonomous research actions by the agent. By default N is 10, though the user can increase the iteration count for more complex analyses. In each iteration, OpenScientist evaluates the query, reviews findings and outputs from any prior iterations, executes new analyses on user-provided datasets, performs targeted searches of the scientific literature, and retrieves and reads relevant abstracts. It then integrates new and prior evidence into an updated set of findings, which are saved in a knowledge state data structure (KSDS) in the form of a JSON file. This iterative loop allows OpenScientist to refine its reasoning, expand its evidence base, and converge toward coherent, evidence-supported conclusions.

At the end of the run, OpenScientist uses the KSDS to produce a final report summarizing all analyses, findings, synthesized knowledge, and evidence-based conclusions. It includes potential follow-up experiments or analyses, when appropriate.

### Availability

OpenScientist is accessible through a public web interface at openscientist.io, which is provided free of charge as resources allow. The web UI supports query submission, data upload, and retrieval of final reports and intermediate outputs. The source code is also available with a permissive Apache 2.0 license at https://github.com/openscientist-io/openscientist for users who prefer to run OpenScientist on their own server infrastructure.

### Case studies

## Results

We evaluated OpenScientist across several biomedical use cases to assess its ability to autonomously analyze data, identify relevant literature, interpret scientific findings, and synthesize coherent conclusions. Case studies were selected to demonstrate OpenScientist’s capability with 1) carrying out a prespecified analysis plan, 2) unspecified data modeling, 3) hypothesis investigation, and 4) hypothesis generation and validation in an external dataset with a negative control (**Table 1**). In addition to the four featured case studies, we evaluated OpenScientist on three further clinical research use cases spanning 5) rare-disease transcriptomics and therapeutic target discovery, 6) placebo-versus-drug adverse-event frequency analysis in curated clinical trial data, and 7) proteomics-based machine-learning interpretation for CSF amyloidosis prediction. For transparency, a full summary of all queries run in OpenScientist during the testing period and the criteria used to select featured case studies is outlined in **Figure 2**. For the featured main text and supplementary case studies, the query and (where possible) the input data, and all output is available in Supplementary Material and on Zenodo (https://doi.org/10.5281/zenodo.18852839).

**Table 1:** Summary of OpenScientist case studies featured in this manuscript.

| Case Study | Purpose | Query | Data | Additional materials | Scientific output |
| --- | --- | --- | --- | --- | --- |
| 1. SEABIRD: Alzheimer's blood biomarkers in a community cohort | Prespecified analysis | Read SEABIRD grant scientific plan and carry out specified analyses. | CSV including amyloid PET, blood biomarkers, and demographics by participant (n=325) | Grant proposal, data dictionaries | Analyzed biomarker performance demonstrating utility of %pTau217 for PET status prediction. |
| 2. Survival prediction by human plasma proteomics | Unspecified modeling | Build a survival prediction model maximizing concordance index on held-out testing set. | CSV of plasma proteomics and demographic data (n=500) | None | Generated survival model utilizing proteomics with improved concordance on held out data and identified most impactful proteins. |
| 3. Neurofibrillary tangle (NFT) transcriptomics in human brain to investigate lysosomal | Hypothesis investigation | Investigate how proteostasis, in particular lysosomal acidification, is rewired by tau pathology. | h5ad file of single cell transcriptomics of neurons with and without NFTs | None | Identified altered expression of transcripts significant to lysosomal acidification in NFTs. |
| acidification |  |  |  |  |  |
| 4. Multiple myeloma (MM) transcriptomics for hypothesis generation and validation | Hypothesis generation | Identify differentially expressed genes between clinical subgroups and hypothesize a biological mechanism for progression of myeloma. | RNASeq data with patients stratified by clinical subgroup (MGUS, SMM, MM) (n=99) | None | Hypothesized UPR failure mechanisms are central to MM progression. |
|  | Hypothesis validation with negative control | Given a set of hypotheses, validate on two datasets. | External RNASeq validation dataset (n=162), scrambled RNASeq dataset | OpenScientist output from initial hypothesis generating run | Appropriately rejected hypotheses in randomized data. |

**Figure 2:**
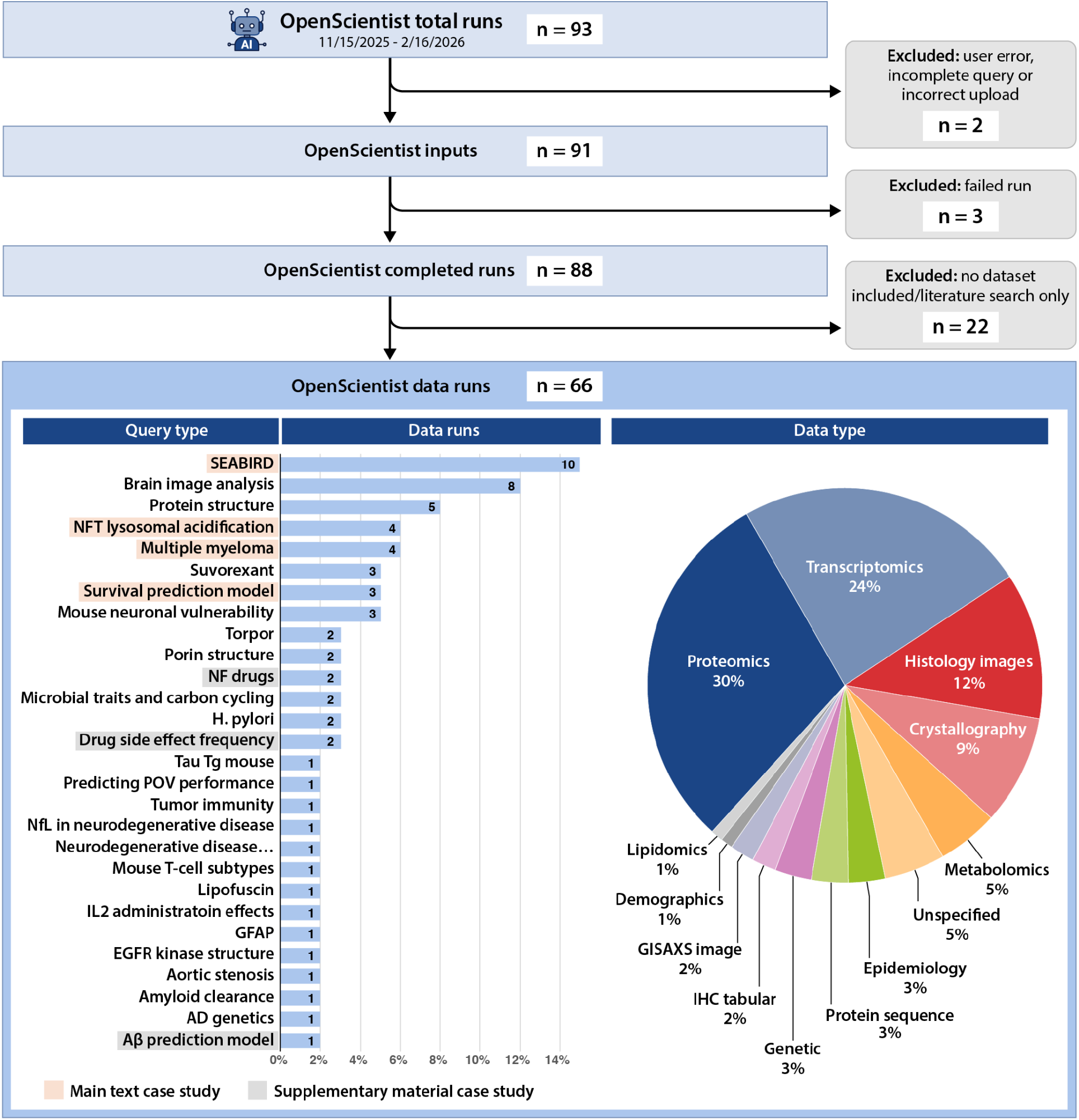
CONSORT diagram outlining all OpenScientist runs during the development period. Two runs failed due to user error. Three runs had technical failures in OpenScientist. Of the remaining 88 OpenScientist runs, 22 did not include a dataset and were limited to a literature search query. In order to emphasize the analytic capability of OpenScientist beyond literature search alone, these were excluded from contention as featured case studies. The 66 remaining runs split into 29 scientific topics, some of which were run multiple times, and covered broad data types. Of the 29 possible topics, 7 are included as case studies, with user engagement and completion of expert evaluation of OpenScientist results being the primary drivers for inclusion. Supplementary materials for the case studies (including input files, where possible, and all OpenScientist outputs) are available on Zenodo (https://doi.org/10.5281/zenodo.18852839)

### Case Study 1: SEABIRD: Alzheimer’s disease blood biomarkers in a community cohort

The first dataset used to evaluate OpenScientist consisted of multimodal clinical, plasma biomarker, and imaging data from a subset of 325 participants enrolled in the Study to Evaluate Amyloid in Blood and Imaging Related to Dementia (SEABIRD, NCT03899844) who underwent blood collection and confirmatory amyloid PET imaging.^12^ Plasma biomarkers (including Aβ42, Aβ40, pTau181, pTau205, pTau217, and total tau) were enriched via immunoprecipitation and measured using liquid chromatography-tandem mass spectrometry.^13,14^

Accompanying data included demographic variables (age, sex, race), *APOE* genotype, cognitive and functional assessments (AD8, Montreal Cognitive Assessment [MoCA], and Clinical Dementia Rating sum of boxes [CDR-SB]). Amyloid PET Centiloid values provide a quantitative reference standard for cerebral amyloid burden.^15^ This dataset represents a clinically realistic, well-phenotyped cohort suitable for benchmarking OpenScientist’s ability to integrate heterogeneous biomedical data and investigate relationships between blood biomarkers, cognition, genetic risk, and imaging-defined pathology.

The prompt for OpenScientist included the raw data in tabular format, data dictionaries, and the grant application for the study, which included relevant background information, the study design, aims, the analytical plans. The query instructed OpenScientist to use the proposal to conduct a full analysis of the data based on the aims and goals of the studies (**Figure 3a**). To address the query, OpenScientist first performed exploratory data analysis to characterize cohort composition, missing data patterns, and the distributions of plasma biomarkers, demographic variables, and cognitive measures. OpenScientist reconstructed the intended analytical pipeline from the grant application, implementing receiver operating characteristic (ROC) analyses to evaluate the discriminative performance of individual plasma biomarkers for amyloid PET status. The system generated executable Python code to reproduce these previously unpublished analyses, including data preprocessing, feature selection, and ROC curve estimation. Exhaustive verification of each generated script and output demonstrated 100% concordance between the scripts and reported results.

**Figure 3:**
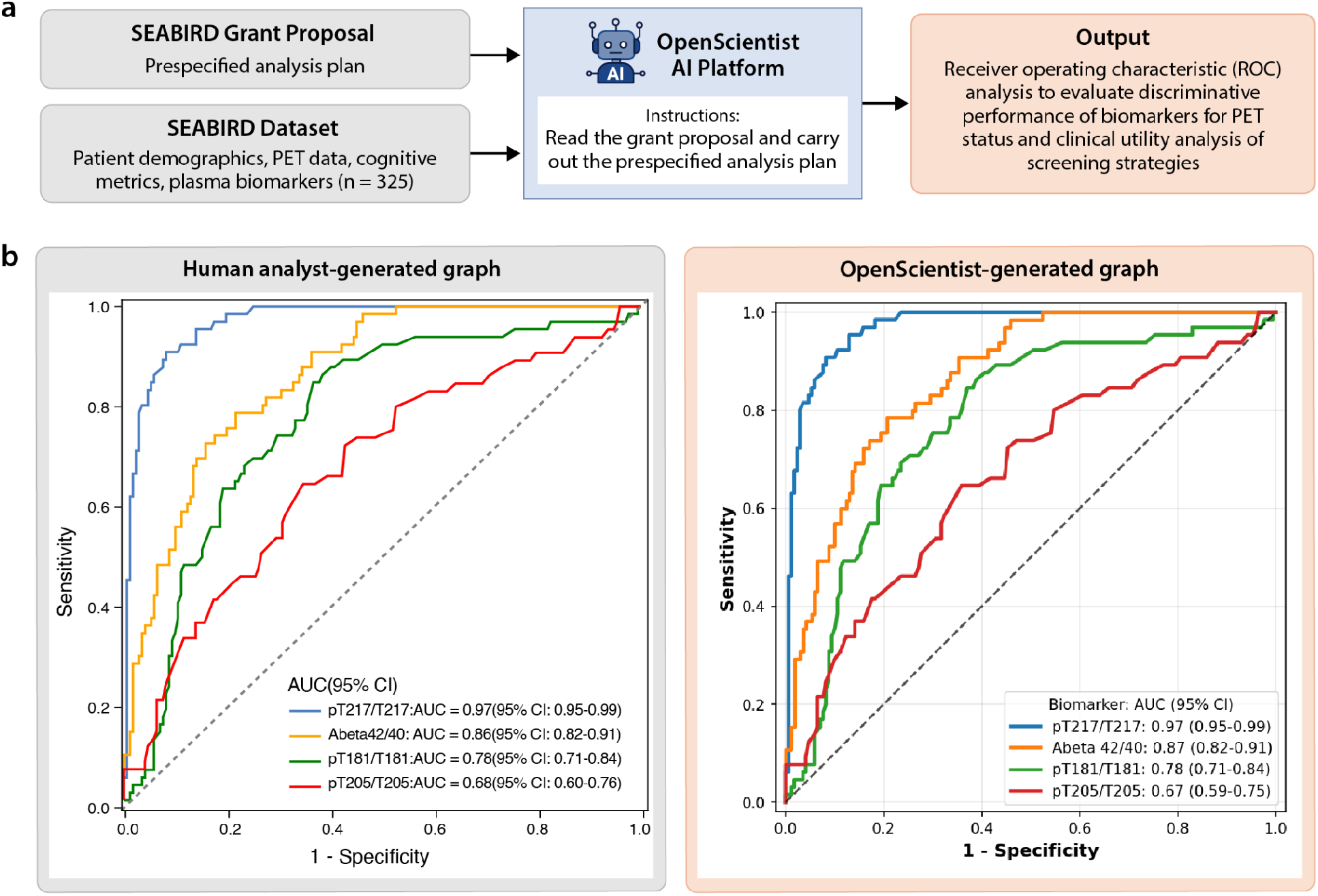
**a)** Workflow: Case Study 1 utilized the SEABIRD dataset which included plasma biomarkers of Alzheimer’s disease, PET Centiloid values, cognitive scores, and patient demographics from a community cohort. OpenScientist was provided the SEABIRD grant proposal and dataset and instructed to carry out the specified analysis plan which included ROC analysis to evaluate the ability of plasma biomarkers for discriminating PET status. **b)** Comparison of human generated analysis of the dataset (left) and OpenScientist generated analysis (right) showing equivalent conclusions.

Consistent with independent human-led analyses of the SEABIRD cohort using Statistical Analysis System (SAS), OpenScientist identified plasma %pTau217 as the highest-performing individual biomarker for distinguishing amyloid-positive from amyloid-negative participants (**Figure 3b**).^16^ In both the human-generated and OpenScientist-generated analyses, %pTau217 substantially outperformed %pTau181, %pTau205, and Aβ42:40, with ROC curves demonstrating superior sensitivity at low false-positive rates. Absolute AUC values differed very minimally between analyses, likely reflecting rounding differences.

During this early exploration phase, we executed the SEABIRD case study in OpenScientist multiple times (n = 10) while iteratively refining our use of the interface. Across these independent runs, we identified several discrepancies in OpenScientist’s performance that flagged inaccuracies in data handling, statistical reporting, and cohort definition. Each discrepancy was traceable to either ambiguity in prompt specification or assumptions made during data ingestion and preprocessing. For example, OpenScientist initially treated empty cells in CSV files as zero values rather than missing data, resulting in misclassification of participants with absent amyloid PET data as amyloid negative. In other instances, the system failed to consistently report sample sizes, confidence intervals, or statistical methods, used a literature-derived Centiloid cutoff rather than the prespecified study threshold, and generalized findings to the full SEABIRD cohort despite the dataset representing only the confirmatory PET subset. Additionally, duplicate records present in the initial data upload were erroneously treated as independent observations. Through iterative prompt refinement and data curation, these issues were resolved by explicitly specifying missing-data handling rules, required reporting standards, cohort boundaries, and the MRI-free Centiloid cutoff of 18.41 for amyloid positivity, as well as by uploading a deduplicated dataset.^17^ While the data represented in **Figure 3** are derived from the final optimized run, a detailed table summarizing the discrepancies observed across runs and the corrective prompt or data modifications implemented is provided in Supplementary Material.

This iterative process highlights a central lesson of agentic AI deployment in biomedical research: reliability depends not only on the model’s analytic capacity, but also on human scientific expertise and oversight to review and validate these systems with explicit instructions, transparent preprocessing logic, and structured validation. Systematic re-running and auditing of analyses were essential for identifying hidden assumptions and ensuring analytic fidelity prior to formal reporting.

### Case Study 2: Survival prediction by human plasma proteomics

The second evaluation dataset consisted of plasma proteomic and demographic data from 500 participants spanning ages 18 to 99 years generated for the Biomarkers of Aging benchmarking challenge.^18^ Proteomic measurements were generated using two Alamar NULISA panels targeting central nervous system and inflammation-related proteins. Accompanying metadata includes demographic variables such as age and sex, as well as survival outcomes defined by time to death. This dataset represents a well-characterized, moderately sized cohort suitable for benchmarking survival modeling approaches and assessing the added value of high-dimensional proteomic features beyond basic demographics.

OpenScientist was prompted with a clearly defined, goal-oriented task: to build a survival prediction model maximizing concordance index on a held-out evaluation set (**Figure 4a**). The instructions specified a reproducible 50%:50% training:testing split, inclusion of all proteomic and metadata features, and comparison against a baseline model restricted to age and sex. The query allowed flexibility in model choice, including traditional Cox proportional hazards models and modern machine learning approaches such as penalized Cox or random survival forests, with explicit encouragement to tune or regularize models to improve performance. This design closely mirrors real-world applied data science workflows and aligns with the official evaluation criteria of the benchmarking challenge.

**Figure 4.**
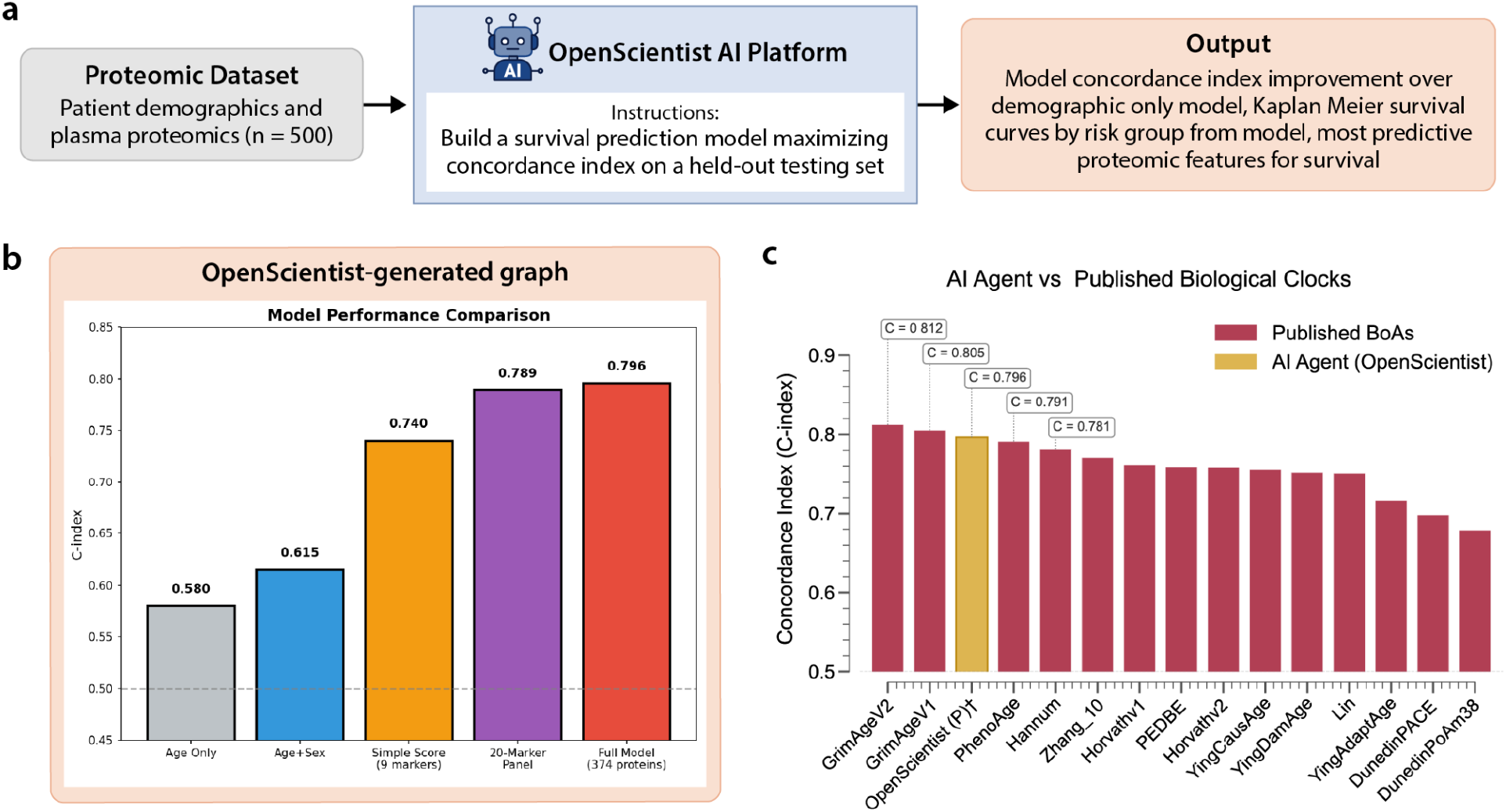
**a)** Workflow: Case Study 2 utilized a dataset including plasma proteomics and demographic data. OpenScientist was prompted to build a survival model. **b)** OpenScientist output graph comparing model performance and identifying proteomic features predictive of survival. **c)** OpenScientist shows comparable performance by c-index compared to published Biomarkers of Aging (BoA) models. † Internal validation (50/50 train-test split); not evaluated on competition holdout set.

OpenScientist’s primary finding was a substantial and consistent improvement in survival prediction when proteomic features were incorporated (**Figure 4b**). The full proteomic model achieved a c-index of 0.796 on the evaluation set, compared to 0.615 for the age and sex baseline model, representing an absolute improvement of 0.181 and a relative improvement of approximately 30%. This magnitude of gain is large for survival modeling and indicates meaningful additional prognostic information contained in the proteomic data. The robustness of this result was supported through multiple validation strategies. Five-fold cross-validation yielded comparable performance estimates, with mean c-indices of 0.811 for the proteomic model and 0.610 for the baseline, and a statistically significant paired comparison. Bootstrap resampling across 100 random splits further demonstrated that the proteomic model outperformed the demographic baseline in all iterations, with a mean improvement of 0.200 and a narrow confidence interval. These analyses reinforce the stability of the performance advantage and reduce concerns about overfitting or favorable data partitioning.

In addition to the full model, OpenScientist derived a reduced nine-marker risk score that retained strong predictive performance, achieving a c-index of 0.740 and stratifying individuals into risk groups with an 8.6-fold difference between lowest and highest risk. This parsimonious model highlights potential translational value and demonstrates that much of the predictive signal can be captured with a limited subset of markers.

From a data handling perspective, OpenScientist correctly merged the two proteomic panels with metadata and appropriately identified predictors and survival outcomes. The use of paired statistical tests to compare model performance was methodologically sound, given that models were evaluated on identical data splits. Visualization outputs, including survival curves and risk score relationships with age, were informative and aligned with best practices for survival analysis, although the absence of ROC or time-dependent AUC plots was noted. Some technical issues occurred in early iterations, including timeouts during elastic net fitting and a variable scoping error when fitting the baseline Cox model. These failures did not materially affect the final conclusions but highlight practical limitations related to execution constraints rather than conceptual flaws in the analytical approach.

OpenScientist identified biologically coherent mortality predictors clustering into distinct pathways: inflammaging (IL-6, IL-33, SPP1/osteopontin, CALCA), neurodegeneration (NEFL, NGF), organ damage (HAVCR1/KIM-1), and immune dysregulation (CD274/PD-L1). Protective factors included markers of T-cell function (CD3E, GZMA) and neural health (CNTN2, ACHE). Key predictors such as NEFL, SPP1, and HAVCR1 were independently supported by prior literature linking them to aging, systemic disease burden, and mortality. Notably, OpenScientist surfaced one highly relevant publication on plasma NEFL and all-cause mortality that had not been identified in the manual review.^19^ On the other hand, several key publications linking top-ranked markers to longevity were missed, including recent work implicating CX3CL1 in human longevity, which was not indexed in PubMed at the time of analysis.^20^ This limitation highlights a dependency on literature source coverage rather than analytical reasoning and underscores the importance of complementary literature retrieval strategies when interpreting AI-generated mechanistic narratives.

Some mechanistic interpretations proposed by OpenScientist extended beyond the current consensus in aging biology. In particular, directional links between immune checkpoint signaling and core aging hallmarks were presented with more confidence than is currently supported by the literature. Additionally, the relative importance of certain predictors, such as CNTN2, was somewhat overstated compared to their ranking in the primary statistical models. These issues reflect challenges in balancing hypothesis generation with evidentiary caution and emphasize the need for human oversight in interpreting mechanistic narratives.

Overall, OpenScientist demonstrated strong performance across key evaluation criteria. The analyses were largely correct, comprehensive, verifiable, and explainable, and the system reached conclusions consistent with expert manual analyses while providing additional robustness metrics and biological insights. According to expert review, “OpenScientist’s model performance was comparable with the top models submitted by participants in the benchmarking challenge using the same data.”^18^ When benchmarked against published Biomarkers of Aging clocks on the same dataset, the OpenScientist model ranked third overall by concordance index, placing it among the top-performing approaches.^21–34^ In terms of efficiency, OpenScientist replicated and extended a complex survival modeling workflow in substantially less time than a human-led analysis, at minimal marginal cost. The primary value lies in rapid model development, systematic validation, and literature synthesis, with manageable risks related to overinterpretation rather than statistical error.

### Case Study 3: Neurofibrillary tangle transcriptomics in human brain to investigate lysosomal acidification

The third evaluation dataset consisted of single-soma transcriptomic data from post-mortem human brain tissue, originally published by Otero-Garcia et al. (2022).^35^ This dataset enables direct comparison of transcriptional programs between neurons harboring neurofibrillary tangles and matched tangle-free neurons from the same donors, providing a unique view of cellular responses to tau pathology. The dataset was obtained from CellxGene and subsetted to two populations of excitatory neurons (Ex1 and Ex2) for analysis.

OpenScientist was prompted with a hypothesis-driven query: to investigate how the proteostasis network is rewired with the appearance of tau pathology, with particular focus on lysosomal acidification. The goal was to test OpenScientist’s ability to build a mechanistic model from an open-ended biological question. Importantly, this query intentionally explored a biological direction beyond the scope of the original analysis. We selected this topic because it was not systematically examined in the original publication, except for a mention of “autophagy” (one of the components of the proteostasis network). By focusing on lysosomal acidification, the query required OpenScientist to interrogate the dataset in a novel direction rather than reproduce prior findings. Our prompt specified a single biological system (proteostasis) and a specific subprocess of interest (lysosomal acidification), but left the analytical approach, pathway and gene selection, and interpretation entirely to the agent.

After data loading and initial exploration, OpenScientist assembled a list of genes largely covering the various arms of the proteostasis network (autophagy, lysosomal, vacuolar-ATPase - vATPase subunits, chaperones, proteasome subunits, and ubiquitin and related genes) to investigate.^36,37^ Amongst those, OpenScientist identified that several subunits of the lysosomal vATPase (the main proton pump responsible for acidification) were significantly upregulated. This observation led to literature searches to understand the role of vATPase subunits in lysosomal function and neurodegeneration as well as links with tau clearance and aggregation. Through several iterations, OpenScientist proposed a “fundamental discovery”: lysosomal acidification is not impaired through impaired vATPase function but through a uniform downregulation of other lysosomal channels (such as MCOLN1-3, TMEM175, CLCN7, or TPC1/2) (**Figure 5**).

**Figure 5:**
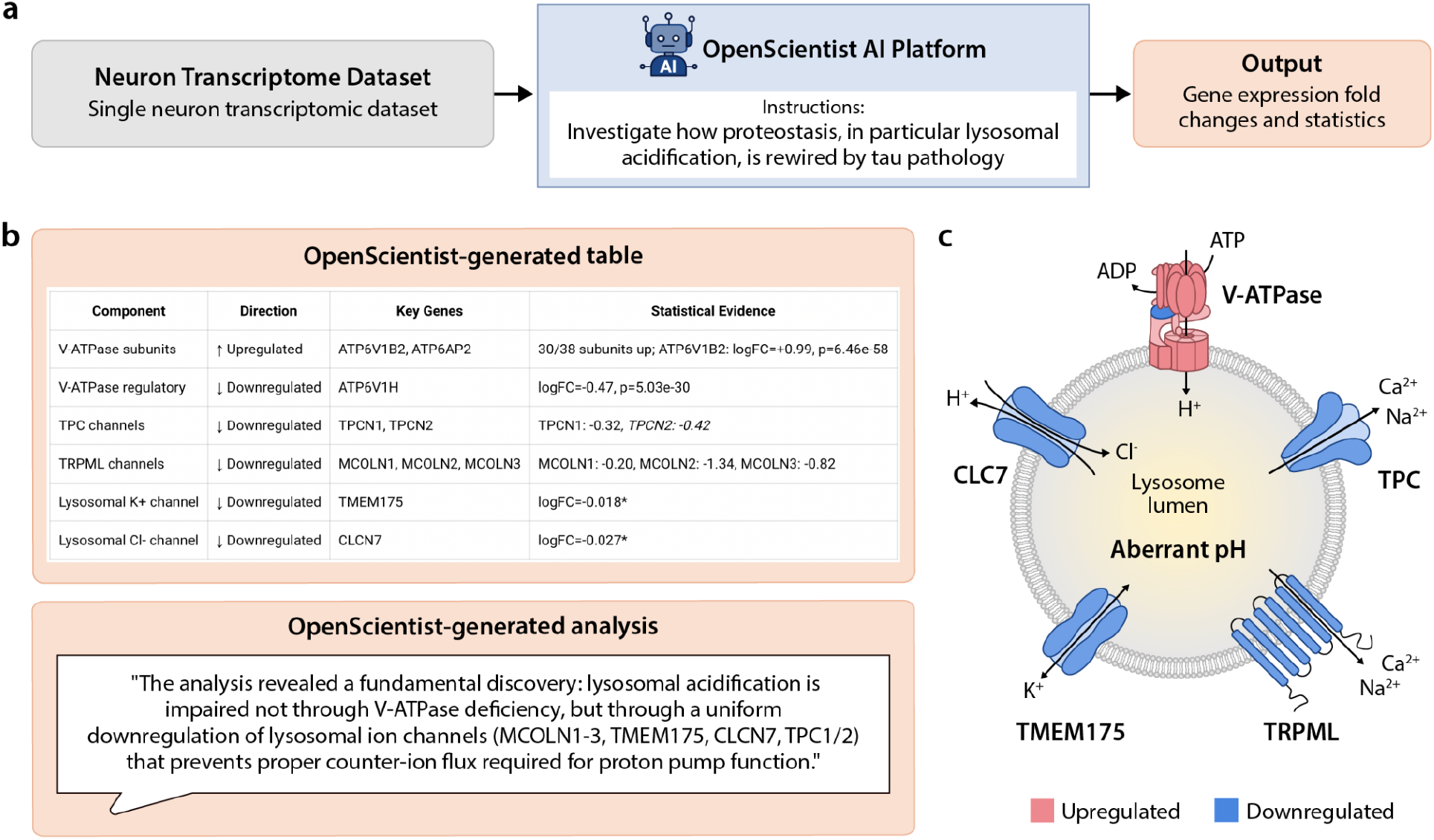
**a)** Workflow: Data included single-soma transcriptomics from neurons with and without neurofibrillary tangles (NFTs). OpenScientist was instructed to investigate the relationship between proteostasis and lysosomal acidification. **b-c)** OpenScientist analyzed gene expression changes and generated a discovery highlighting a relationship between neurons with NFTs and impaired lysosomal acidification via downregulation of lysosomal ion channels that was verified by a human expert.

Overall, the main findings were independently verifiable (i.e. r=0.983 correlation between claimed and verified log2 fold changes when computed by a domain expert). The statistical approach was standard but did not account for the hierarchical structure of the data (8 donors, multiple cells per donor). OpenScientist noted this limitation and assessed cross-donor consistency, but did not suggest more sophisticated approaches (e.g. mixed-effect models). The proposed hypothesis is consistent with current knowledge about mechanisms controlling lysosomal pH, in particular the imbalance between H^+^ influx via the vATPase and H^+^ leak via cation channels such as TMEM175 or potentially MCOLN1 (TRPML1).^38^

This analysis was not without limitations. First, in terms of scope, despite an initial large-scale investigation, OpenScientist mostly focused on lysosomal acidification (which was only part of the prompt) and autophagy without investigating other aspects of the proteostasis network. This limitation could probably be overcome by refining the prompt or adopting the scientist-in-the-loop approach allowing feedback during the analysis, which was added as a feature (co-investigate mode). Additionally, some complex biological pathway interpretations relied heavily on a small number of significant genes demonstrating an overly reductionist approach. Last, OpenScientist demonstrated a tendency to over-rely on statistical significance over effect size (e.g. TMEM175 was included in the final model despite a reported Log2 fold change of -0.018 because it reached statistical significance).

In conclusion, OpenScientist demonstrated strong performance across all criteria and allowed conclusions comparable to several days of expert work to be reached within 10 iterations. Despite the limitations listed above, OpenScientist proposed a relevant mechanism, aligning well with current literature, and going beyond the initial prompt. This case highlights the relevance of AI co-scientists such as OpenScientist to extract novel knowledge from latent data (published, well-curated datasets harboring analytical potential that exceeds the scope of their original investigations).

### Case Study 4: Multiple myeloma transcriptomics for hypothesis generation and validation

#### Phase 1: Initial discovery and hypothesis generation

The fourth case study used multiple myeloma (MM) gene expression data and tested OpenScientist’s ability to: (1) generate novel mechanistic hypotheses from RNA-seq data (n=99 samples, 33,297 genes) spanning the myeloma progression from normal plasma cells to monoclonal gammopathy of uncertain significance (MGUS) to smoldering multiple myeloma (SMM) to active MM; (2) independently validate these hypotheses using an external cohort (Mayo Clinic, n=162 samples, GSE6477); and (3) correctly identify when validation fails using randomized negative controls (**Figure 6a**). This approach directly addresses the critical question of whether autonomous AI systems can perform reliable scientific validation without human supervision.

**Figure 6:**
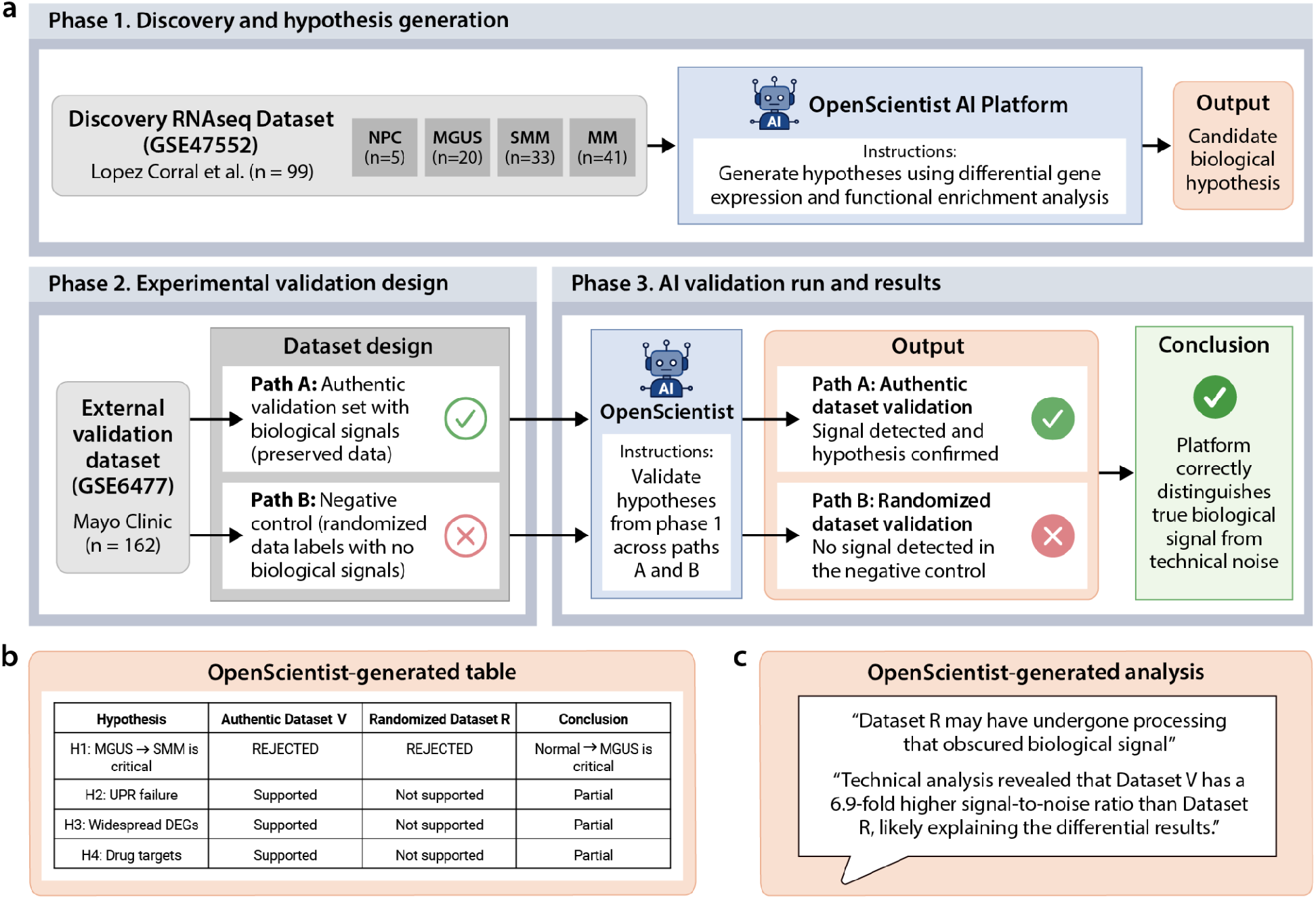
Schematic of the AI-driven hypothesis generation and rigorous validation framework. The study began with the OpenScientist platform analyzing the multiple myeloma dataset (n=99) to identify differentially expressed genes and functional pathways across the multiple myeloma progression spectrum (NPC normal plasma cell, MGUS monoclonal gammopathy of uncertain significance, SMM smoldering multiple myeloma, MM multiple myeloma). To validate these findings, a dual-pronged approach was employed using the dataset (n=162). A negative control was established by randomizing disease labels (Dataset R) in a copy of the validation set (Dataset V). OpenScientist successfully confirmed hypotheses in the authentic data while correctly identifying the absence of reproducible signals in the randomized control, confirming the platform’s analytical specificity.

OpenScientist’s initial analysis of the López-Corral et al. (2014) dataset identified 19,438 differentially expressed genes (58.4% of genome, FDR<0.05) organized into four distinct transcriptional programs.^39^ The platform proposed a novel unfolded protein response (UPR) failure model, positing that MM progression involves three phases: compensated endoplasmic reticulum (ER) stress at MGUS (218 genes showing +16.3% upregulation), catastrophic UPR collapse at the MGUS to SMM transition (81.4% of gene changes), and proteotoxic crisis in active MM. Critically, OpenScientist identified a regulatory balance shift where the ratio of upregulated to downregulated genes crosses parity between SMM and MM, suggesting a “point of no return” in malignant transformation.

### Phase 2: Experimental validation design

To rigorously evaluate OpenScientist’s capacity for autonomous scientific validation, we implemented a dual-pronged approach using two independent multiple myeloma gene expression datasets (Dataset V, n=162; Dataset R, n=162). Both cohorts contained samples spanning the complete disease spectrum from normal plasma cells through MGUS, SMM, newly diagnosed MM, and relapsed MM. Critically, Dataset R represented a negative control: we randomized disease stage labels while preserving all other data characteristics, effectively eliminating biological signal while maintaining technical structure. This design enabled us to test whether OpenScientist could discriminate true biological validation (Dataset V) from spurious patterns in randomized data (Dataset R), a fundamental requirement for autonomous scientific reasoning.

OpenScientist was provided with four hypotheses derived from its initial López-Corral dataset analysis: (H1) the MGUS to SMM transition represents the critical “point of no return” in disease progression, (H2) UPR failure mechanisms drive malignant transformation, (H3) widespread differential gene expression characterizes disease transitions, and (H4) specific drug targets can be identified from expression profiles. The platform was tasked with independently validating these hypotheses using both datasets without prior knowledge of the randomization manipulation.

### Phase 3: AI validation results

OpenScientist’s performance demonstrated sophisticated capacity for evidence-based scientific reasoning. Most notably, the platform correctly rejected Hypothesis 1 in both datasets based on empirical observations: the Normal to MGUS transition showed substantially larger transcriptional reprogramming (615 DEGs in Dataset V, 112 DEGs in Dataset R) compared to the MGUS to SMM transition (268 and 27 DEGs respectively). This rejection occurred despite H1 being OpenScientist’s original prediction, indicating the platform prioritizes empirical evidence over self-consistency, a critical attribute for scientific discovery systems. A copy of the OpenScientist’s hypothesis summary results can be found in **Figure 6b**. The rejection was appropriately replicated across both datasets, with Normal to MGUS consistently ranking as the largest transcriptional change regardless of technical platform or data quality. This finding represents a genuine biological insight: the initial transformation to the precursor MGUS state, rather than progression to overt malignancy, may constitute the major molecular inflection point in MM pathogenesis. The willingness to reject an original hypothesis when confronted with contradictory evidence demonstrates that OpenScientist employs genuine hypothesis testing rather than confirmation bias.

OpenScientist’s validation outcomes diverged appropriately between the two datasets, providing strong evidence for signal discrimination capacity. In Dataset V (true biological data), three hypotheses were supported: H2 (UPR failure mechanism with XBP1 ρ=-0.216, p=0.006; BiP ρ=-0.253, p=0.001), H3 (6,653 total DEGs across all pairwise comparisons), and H4 (BCMA log_2_FC=+0.92, p=0.0006; PSMB5 log_2_FC=+0.84, p<0.0001). In stark contrast, Dataset R (randomized labels) showed no support for these hypotheses: UPR genes showed no significant changes (all p>0.2), only 582 total DEGs were identified, and differential expression analysis failed to reach statistical significance for key drug targets. This differential validation performance demonstrates appropriate statistical stringency. The platform did not simply validate hypotheses indiscriminately; rather, it applied consistent statistical criteria (FDR correction, effect size thresholds) that naturally rejected spurious patterns in the randomized dataset while detecting genuine signals in the true data. The contrast is particularly striking for H3 (widespread differential expression): 100% of pairwise comparisons in Dataset V showed >50 DEGs, compared to only 40% in Dataset R, a quantitative discrimination of signal quality.

Perhaps most encouraging for deployment of autonomous discovery platforms, OpenScientist independently identified fundamental problems with Dataset R without explicit knowledge of the randomization manipulation. The platform reported that Dataset R exhibited a 6.9-fold lower signal-to-noise ratio compared to Dataset V and flagged concerns about data processing that may have “obscured biological signal.” While OpenScientist did not explicitly deduce that labels had been randomized, it correctly recognized that the dataset exhibited characteristics incompatible with expected biological patterns, namely, drastically reduced differential expression signal and inconsistent biomarker progression trends.

This capacity for self-assessment addresses a critical vulnerability in AI-driven science: the “garbage in, garbage out” problem. Rather than generating confident but meaningless results from compromised data, OpenScientist exhibited appropriate epistemic humility by acknowledging dataset limitations and qualifying its conclusions accordingly. The platform noted that “biomarker progression trends showed opposite directions between datasets” and explicitly recommended follow-up experiments including “re-analyze Dataset R with alternative normalization to recover biological signal.” This represents a form of autonomous quality control, where the system recognizes when input data quality undermines inference validity.

## Discussion

We present OpenScientist, an open-source agentic AI platform designed to support semi-autonomous scientific discovery through transparent, iterative analysis and reasoning steps. Across multiple clinical biomedical use cases, OpenScientist demonstrated the ability to integrate user-provided data with published literature, perform bespoke computational analyses, and synthesize mechanistic insights into coherent, verifiable results and testable hypotheses. Together, these results indicate that agentic AI systems can meaningfully augment scientific workflows when they are built with auditability, modularity, and interpretability as core design principles. In particular, our findings suggest that such systems may be especially valuable for accelerating clinical studies, where investigators must rapidly integrate heterogeneous human datasets, extract actionable patterns, and prioritize follow-up analyses under constraints of time, cost, and domain expertise.

OpenScientist independently, without prior knowledge of the results of human scientist analyses, identified novel discoveries in four clinical case studies: (1) a prespecified analysis in a community-based Alzheimer’s disease biomarker cohort, (2) unsupervised modeling for plasma proteomic survival prediction, (3) hypothesis investigation in single-cell transcriptomic data from neurons with neurofibrillary tangles, and (4) hypothesis generation with validation in a multiple myeloma dataset with a randomized negative control. The results and findings were remarkably similar to what human expert scientific teams determined independently, with a few important differences.

From a practical standpoint, OpenScientist improves research efficiency by automating labor-intensive components of scientific workflows, including literature synthesis, data analysis, and iterative hypothesis refinement. By integrating these steps within a single auditable pipeline, the platform reduces time and cost expense while preserving human oversight. In the use cases presented here, the typical cost per OpenScientist run is currently less than $10, with domain scientists estimating data analysis time savings on the order of weeks to months per analysis. Support for multiple LLM providers with integrated cost tracking further enables deployment within predefined resource and budget constraints.

A central contribution of OpenScientist is its open-source and fully auditable architecture, which addresses key limitations of existing AI scientist platforms. While many recent systems demonstrate impressive capabilities, they remain proprietary, limiting reproducibility, customization, and independent validation. In contrast, OpenScientist exposes its full hypothesis generation, analysis, and reasoning pipeline, enabling researchers to inspect intermediate steps, validate outputs, and modify workflows to suit domain-specific needs. Further, the transparent nature supports explainable AI, critical review of performance, and open expert scientist testing and refinement of the rapidly evolving AI for science field. This transparency is essential for building trust in AI-assisted discovery and for aligning autonomous systems with established scientific standards.

OpenScientist’s modular skills architecture further enhances extensibility and adaptability. By separating domain-agnostic reasoning components from domain-specific analytical skills, the platform allows researchers to extend functionality without altering core logic. This design supports community-driven development of specialized skills tailored to particular datasets, experimental paradigms, or biomedical domains, broadening OpenScientist’s applicability while preserving a stable and interpretable core.

Across the use cases presented here, OpenScientist demonstrated strong but imperfect performance. We noted several modes of failure in OpenScientist, including (i) statistical reliability problems (e.g., lack of cross-iteration multiple-testing correction and inconsistent hypothesis prioritization formulas), (ii) domain-science errors in some skills (for example, incorrect biochemical interpretation of metabolite ratios as flux proxies or inappropriate statistical treatment of RNA-seq count data), and (iii) architectural limitations in the reporting and literature review pipeline that can compound summarization errors (e.g., the final report being generated from summaries rather than directly interrogating the data, and literature summaries lacking transparent citation tracking). Potential remediation approaches include updating Agent Skills, introducing explicit statistical safeguards (e.g., FDR or family-wise error tracking across iterations), enforcing consistency checks across skills and scoring formulas, adding domain-aware validation layers for analysis code, and restructuring the report and literature synthesis stages so the agent can directly access data, provenance, and cited sources when generating conclusions. (iv) limitations revealed through systematic validation, including incomplete rejection of hypotheses in negative-control and randomized analyses. While the system reliably reproduced known analytical results, implemented appropriate statistical workflows, identified biologically coherent signals, and generated plausible, testable hypotheses, validation experiments showed that it does not yet consistently achieve complete hypothesis rejection when underlying biological structure is removed. In randomized control analyses, a subset of hypotheses remained statistically supported despite elimination of true signal, indicating incomplete discrimination between genuine biological effects and residual structure. These are similar to modes of failure and mistakes made by human scientists. Similarly, continued training and refinement are likely to improve the expertise, accuracy, and impact of the system’s analyses.

Our findings suggest that current autonomous scientific AI systems, including OpenScientist, should be viewed as powerful accelerators of hypothesis testing and analysis, rather than fully independent scientific actors. Although OpenScientist demonstrates sophisticated reasoning, statistical execution, and literature integration, its susceptibility to partial validation under adversarial conditions highlights the continued need for expert scientist oversight, particularly for mechanistic interpretation and causal inference.

Based on these observations, we identify three immediate priorities for advancing autonomous scientific AI. First, validation frameworks must be strengthened to ensure robust and consistent development, testing, transparency, and use of scientific AI. For example, scientific AI reports should meet the same standards as scientific reports, including controlling for multiple comparisons with full accounting of all *in silico* experiments with pre-specified vs. post-hoc findings clearly identified, full disclosure and reporting of both positive and negative findings, and development and testing by expert scientists. Second, autonomous reasoning systems should be integrated with experimental validation, such as automated design of perturbation or intervention experiments to directly test causal claims. Third, prospective clinical and translational studies are needed to determine whether AI-generated predictions improve real-world patient outcomes. Only through such rigorous evaluation can the field determine whether platforms like OpenScientist represent transformative advances in biomedical discovery or sophisticated pattern-recognition systems that still require extensive human supervision.

For sensitive clinical datasets and unpublished analyses, OpenScientist can be deployed on-premises so data handling remains within existing institutional governance. However, full data containment would minimally require that both the OpenScientist runtime and the underlying LLM be hosted within the same secure environment; use of external model APIs may result in transmission of the user’s input data. Similarly, intellectual property concerns (e.g., unpublished hypotheses or analysis outputs) may require self-hosted deployments. Further, since agentic systems can execute arbitrary code, there still remains a risk of the execution of arbitrary code that could destroy or exfiltrate sensitive data.

Taken together, these results support OpenScientist as a useful and still maturing co-scientist. Its open-source design enables customizable deployment at any institution, including within institutionally controlled environments, allowing sensitive clinical and genomic data to remain within existing governance frameworks while supporting rigorous benchmarking and adversarial testing. By making both successes and limitations explicit, OpenScientist serves not only as a functional discovery platform but also as a testbed for defining standards of validation, transparency, and responsibility in autonomous scientific research. Because the use cases presented here all involved human clinical datasets, OpenScientist may be particularly well positioned to support the acceleration of clinical studies by shortening the path from complex data to interpretable findings, candidate biomarkers, and testable translational hypotheses. Continued development focused on validation rigor, experimental grounding, and prospective outcome testing will be essential to optimize such systems to be deployed with confidence in high-stakes biomedical settings.

## Data Availability

All data produced in the present work are contained in the manuscript and supplemental information.

https://doi.org/10.5281/zenodo.18852839

## Acknowledgements

This work was organized through and supported by C-BRAIN (c-brain.org). This work was also supported by resources and effort provided by the Tracy Family Stable Isotope Labeling Quantitation Center (Principal Investigator (PI) RJB) established by the Tracy Family, Richard Frimel & Gary Werths, GHR Foundation, Pat and Jane Tracy, Anonymous, Anne & Ray Capestrain, Community Foundation Serving West Central Illinois and Northeast Missouri, JTL Family Fund, Payne Family, Mary & Jay Sullivan, Tracy Family Foundation, Catherine & Tom Tracy, Community Foundation for the Land of Lincoln, Jim & Jil Tracy, Joe & Jill Tracy, Sonja & Robert M. Willman, Boniface Foundation, Jean & Michael Buckley, Ann Liberman, Clemence S. Lieber Foundation, Mary Schoolman & Dr. James Hinrichs, and Susan & Scott Stamerjohn brought together by The Foundation for Barnes-Jewish Hospital. SEABIRD was funded through NIA RF1AG061900 (PI: RJB). JHC, NLH, CJM and JTR were supported by the NIH Monarch Initiative 2R24OD011883 and by the Director, Office of Science, Office of Basic Energy Sciences of the U.S. Department of Energy Contract No.

DE-AC02-05CH11231. We thank Drs. Suzanne Schindler, Siobhan Sutcliffe, Ganesh Babula, and Eric Lenze for their guidance of the SEABIRD study.

## Competing Interests

RJB has received research funding from Avid Radiopharmaceuticals, Janssen, Roche/Genentech, Eli Lilly, Eisai, Biogen, AbbVie, Bristol Myers Squibb, and Novartis. Washington University and RJB have equity ownership interest in C2N Diagnostics. Washington University, RJB, VO, JGB, and NRB receive income based on technology (stable isotope labeling kinetics, blood plasma assay, methods of diagnosing AD with phosphorylation changes, neurofilament light chain assays and materials) licensed by Washington University to C2N Diagnostics. RJB receives income from C2N Diagnostics for serving on the scientific advisory board. RJB serves as an unpaid member on a scientific advisory board for Biogen. TLSB has served as a paid and unpaid consultant to Eisai, Siemens, Biogen, Janssen, Eli Lilly, Sora Neuroscience and Bristol-Myers Squibb. All other authors declare no competing interests.

## Notes

### Author Declarations

The datasets used in our study have been de-identified. Washington University in St. Louis Institutional Review Board gave ethical approval for the SEABIRD study (IRB #201902081).

