## Supplemental Text for "OpenScientist: evaluating an open agentic AI co-scientist to accelerate biomedical discovery"

### Supplemental Information

In addition to the four case studies described in the main manuscript, we tested OpenScientist on three additional clinical research questions (Case Studies 5, 6, and 7). The results are described below.

#### Case Study 5: Neurofibromatosis transcriptomics and therapeutic candidate identification

To evaluate OpenScientist's ability to perform exploratory analysis and therapeutic target discovery in rare disease datasets, we tested the agent using bulk RNA sequencing data from patients with neurofibromatosis obtained from multiple institutions and biobanked within the Johns Hopkins NF1 Biorepository.[1,2] The prompt asked OpenScientist to analyze transcriptomic data and identify candidate therapeutic pathways and genes. In order to test its exploratory abilities, no clinical metadata was provided to the agent.

Neurofibromatosis is a tumor-predisposition syndrome characterized by diverse tumor types that develop throughout a patient's lifetime. While many tumors are benign, malignant peripheral nerve sheath tumors (MPNSTs) represent an aggressive manifestation of the disease and have limited treatment options. As a result, identifying new therapeutic targets remains an active area of investigation. The dataset provided to OpenScientist represented a realistic rare disease research scenario. Data were aggregated across institutions and exhibited substantial batch effects that required correction before meaningful downstream analysis.

OpenScientist successfully loaded and explored the transcriptomic datasets and generated a structured report describing its analytical workflow and candidate therapeutic targets. Despite the absence of clinical labels, the agent identified three major tumor clusters based on gene expression patterns. One cluster was inferred to represent MPNST samples based on expression signatures associated with malignant nerve sheath tumors and known oncogenic signaling pathways. OpenScientist also detected the presence of batch effects across datasets and applied mitigation strategies before performing downstream analyses.

The agent identified several targetable genes and pathways supported by the data. Notably, OpenScientist recovered well-established signaling pathways in neurofibromatosis biology, including MEK1/MAP2K1, CDK4, and mTOR signaling.[3–8] The recovery of these known targets provides a useful internal validation of the analysis. In addition to these established pathways, OpenScientist proposed several less characterized candidates. One candidate, IGF2BP1, has been reported in a recent preprint, while additional targets including ULBP1/ULBP2 and the mitotic kinase TTK/MPS1 seem novel within the neurofibromatosis literature.[9] The agent also outlined potential next steps for validating these findings, including additional computational analyses and experimental validation strategies.

This case study also highlighted several limitations of the current system. The analytical rigor of the output was sensitive to prompt design. Earlier prompts that contained fewer instructions regarding RNA sequencing analysis and batch effect correction produced less rigorous results. In addition, although OpenScientist performs literature retrieval during analysis, it labeled IGF2BP1 as a novel finding despite the presence of a recent preprint, illustrating challenges in distinguishing emerging literature from truly novel discoveries.

Overall, this case study demonstrates that OpenScientist can perform exploratory analysis of heterogeneous transcriptomic datasets in a rare disease context, identify known disease pathways, and generate plausible new therapeutic hypotheses. At the same time, the results emphasize the importance of carefully structured prompts and expert review when interpreting discoveries generated by agentic AI systems.

#### Case Study 6: Drug and placebo side effect frequencies in a curated clinical trial dataset

A persistent challenge in drug safety is that adverse-event frequencies observed in trials reflect both pharmacology and non-pharmacologic contributions. Placebo groups in clinical trials often exhibit substantial adverse-event reporting, which can arise from baseline symptom burden, expectations and attribution processes (nocebo effects), and differences in how adverse events are elicited and recorded across trials.[10] These placebo-attributed adverse effects can contribute to treatment discontinuation and poor adherence and can bias the interpretation of drug safety profiles if placebo baselines are ignored.[11] Recent large-scale syntheses in specific settings (for example vaccine trials) further illustrate that a large fraction of reported systemic symptoms can occur in placebo arms, underscoring the scale and generality of this problem.[12]

In addition, computational and chemoinformatic efforts increasingly use drug-label resources to study adverse drug reactions at scale. Drug phenotype and side-effect profiles have been widely used as informative signatures for inferring mechanisms and generating repositioning hypotheses, demonstrating that systematic analysis of clinical phenotype patterns can yield actionable leads.[13] Motivated by these lines of work, and following the frequency-class extraction and encoding framework described by Galeano et al. using the Side Effect Resource (SIDER)-derived frequency information, we curated a dataset restricted to drug–side effect associations where placebo frequency is equal to or higher than drug frequency.[14] This curated “placebo-dominant” subset is valuable precisely because it challenges default causal intuition (that drug exposure should increase side effects) and supports a different class of scientific questions: which symptoms and drugs most often show placebo-dominant frequencies, how large those differences typically are, and whether the findings persist under perturbations and negative controls. As a result, it serves as a high-signal evaluation task for OpenScientist, testing whether the system can help a scientist rapidly formulate and execute rigorous, audit-ready analyses that meaningfully guide subsequent research decisions.

To evaluate OpenScientist's ability to interrogate the structured clinical dataset that we built, and perform rigorous exploratory analysis under constrained conditions, we tested the agent using a curated Excel dataset of drug-side effect associations derived from clinical trial reporting. The dataset contained four fields: drug_name, side_effects, frequency_drug, and frequency_placebo, with frequencies encoded on a five-level ordinal scale ranging from very rare to very frequent. The data were specifically curated to include only drug-side effect pairs in which placebo frequency was equal to or greater than drug frequency. OpenScientist was instructed to use only the Excel content to perform dataset verification, answer two ranking questions restricted to rows where placebo frequency was strictly higher than drug frequency, and provide at least one robustness check and one negative control.

This case study was designed as a structured test of whether an agentic AI system could accurately characterize a small tabular dataset, answer prespecified analytical questions, and distinguish well-supported findings from unsupported extensions beyond the provided data. Because the prompt explicitly restricted the analysis to the spreadsheet alone, this case also served as a test of whether OpenScientist would remain grounded in the input data rather than introducing external annotations or unsupported inferences. Because this curated subset was assembled for this study and we did not identify a publicly released version of the same file, direct leakage of the exact dataset into OpenScientist is unlikely; however, the underlying source materials (e.g., drug labels) are public.

OpenScientist performed strongly on the core Excel-derived tasks. It correctly reported the major dataset characteristics, including total row count, numbers of unique drugs and side effects, absence of missing values, absence of duplicate rows, and the lack of any violations of the curated placebo-greater-than-or-equal-to-drug rule. It also correctly quantified the two main subsets in the data, with most rows showing equal placebo and drug frequencies and a smaller subset showing placebo strictly higher than drug. Within the placebo-higher-than-drug subset, OpenScientist accurately identified the leading drugs and side effects, including cabozantinib among drugs and headache among side effects. It also correctly described the typical magnitude of the placebo-drug difference, which was usually small, and recovered the main transition patterns across ordinal frequency levels.

Beyond simple ranking, OpenScientist also generated robustness analyses. It reported that the leading drugs and side effects were stable under bootstrap resampling, which was confirmed by independent human audit when assessed using a tie-aware definition of leaderboard inclusion. It also implemented a permutation-based negative control showing that the observed number of placebo-greater-than-drug rows was far lower than expected under random pairing, indicating strong non-random structure in the dataset. Together, these results demonstrate that OpenScientist could move beyond descriptive counting to perform more rigorous checks of stability and non-triviality.

The main limitations emerged when the agent extended beyond what could be supported by the Excel file alone. In particular, OpenScientist reported analyses involving therapeutic class, body system, subjective versus objective symptom categories, and logistic regression using derived variables such as mechanism complexity. These analyses were not computable from the spreadsheet as provided and therefore were not supported by the stated input constraints. In one case, a permutation comparison was directionally interpreted correctly but paired with an inconsistent p-value framing. These issues highlight an important failure mode in which the agent performs well on grounded quantitative tasks but may still overextend into unsupported analytic layers when not tightly constrained.

Overall, this case study shows that OpenScientist can accurately verify curated spreadsheet data, answer prespecified ranking questions, and apply robustness and negative-control analyses in a transparent and largely reliable manner. At the same time, it underscores the importance of strict grounding and expert review, particularly when an agent is asked to remain within the bounds of a limited structured dataset.

| Check / claim | OpenScientist report | Independent Human audit | Verdict | Notes |
|---|---|---|---|---|
| Total rows | 2,247 | 2,247 | Correct | Exact match. |
| Unique drugs | 287 | 287 | Correct | Exact match. |
| Unique side effects | 370 | 370 | Correct | Exact match. |
| Missing values | 0 (all columns) | 0 (all columns) | Correct | Exact match. |
| Exact duplicate rows | 0 | 0 | Correct | Exact match. |
| Curated constraint violations (placebo < drug) | 0 | 0 | Correct | Exact match. |
| Subset sizes | placebo=drug 1,893 (84.2%); placebo>drug 354 (15.8%) | placebo=drug 1,893 (84.2%); placebo>drug 354 (15.8%) | Correct | Exact match. |
| Frequency code “2” rarity | drug=2 occurs 2 times; placebo=2 occurs 0 times | drug=2 occurs 2 times; placebo=2 occurs 0 times | Correct | Confirms code-level sparsity (effective levels mostly 1,3,4,5). |
| Top drugs (placebo>drug), leading entries | cabozantinib 14; olmesartan medoxomil 12; levocarnitine 10; famciclovir/ciclosporin/tamsulosin/carmustine 9 | Same leaders, same counts | Correct | Leaders are correct. |

| | | | | |
|---|---|---|---|---|
| “Top 10” drugs cutoff uniqueness | Presented a top-10 list | Cutoff has ties: 8 drugs are tied at count=6 | Correct with caveat | Not wrong, but should explicitly note ties; ranks 9–10 are not unique unless tie-break rule stated. |
| Top side effects (placebo>drug), leading entries | headache 24; GI pain 14; abdominal pain 14; nausea 12; vomiting 9; etc. | Same leaders, same counts | Correct | Leaders are correct. |
| “Top 10” side effects cutoff uniqueness | Presented a top-10 list | Cutoff has ties: 9 side effects are tied at count=5 | Correct with caveat | Same tie issue at the boundary. |
| Delta distribution in placebo>drug subset | delta=1: 312 (88.1%); delta=2: 14 (4.0%); delta=3: 28 (7.9%) | Same | Correct | Exact match. |
| Main transitions (placebo>drug rows) | 4 to 5: 205; 3 to 4: 105; 1 to 4: 28; 1 to 3: 11; 3 to 5: 3; 2 to 3: 2 | Same | Correct | Exact match. |
| Spearman correlation: frequency_drug vs placebo>drug indicator | About -0.40, p < 0.0001 | -0.4063 (p ~ 4.8e-90) | Correct | Exact P-value was not provided |
| Bootstrap “stability” of top findings | Reports high stability for leaders | Tie-aware bootstrap confirms leaders are very stable (e.g., headache 1.00; cabozantinib ~0.997 top-set inclusion in 300 resamples) | Correct with definition note | Stability depends on definition (top-5 vs top-10; tie-aware vs strict). Core message holds. |
| Permutation baseline: expected placebo>drug under random pairing | Expected placebo>drug ~572.5 (about 25.5%) | 572.4 mean in 300 perms; 2.5–97.5% approx. 555.5–587.0 | Correct | Confirms observed placebo>drug (354) is far below permutation baseline (strong concordance structure). |

| | | | | |
|---|---|---|---|---|
| Permutation: max side effects per drug (observed vs permuted) | Observed 14; perm mean 18.8; reported p=1.000 | Observed 14; perm mean 18.68; 2.5–97.5% 16–22; lower-tail p < 1/300 | Partially correct | The comparison is right, but the p-value direction is inconsistent (p=1.000 corresponds to "is observed unusually high," not "unusually low"). |
| Permutation: max drugs per side effect | Observed 24; slightly above permuted range; p around 0.02 | Observed 24; 2.5–97.5% approx. 14.5–23.0; upper-tail p ~0.0167 (300 perms) | Correct | Confirms one side effect (headache) is unusually "shared" across drugs under placebo>drug vs permutation. |
| Therapeutic class / body-system / subjective-objective analyses | Reported as results | Not computable from Excel columns alone | Not supported by Excel | Requires external mapping/annotation that was not provided. |
| Logistic regression using "mechanism complexity" etc. | Reported metrics (accuracy/AUC, baseline improvement) | Not reproducible from Excel; internal metric logic is questionable given class imbalance | Not supported + inconsistent | Unclear where the data come from and whether it is correct. |

**Supplementary Table 1:** Side-by-side comparison of OpenScientist outputs and independently verified results for the curated placebo-dominant dataset. Columns report the analytical claim, OpenScientist's output, independently verified values by a human curator derived solely from the spreadsheet, an agreement verdict, and clarifying notes. Only Excel-derived quantities were considered in the audit.

#### Case Study 7: Proteomics machine learning model trained to predict CSF Aβ42/40

To evaluate OpenScientist's ability to interpret model-derived biological signals and integrate pathway-level information with prior knowledge, we tested the agent on results from a proteomics-based predictive model of amyloidosis. An elastic net regression model had been trained to predict CSF Aβ42/40 ratios from CSF proteomic measurements. In this model, negative protein weights indicate that higher protein abundance is associated with lower Aβ42/40 ratios, consistent with greater amyloid pathology, while positive weights indicate the opposite relationship. Proteins from the model were subsequently examined for correlation with chronological age in the same CSF proteomics dataset. Proteins showing age-associated abundance changes were then subjected to Reactome pathway enrichment analysis using the Metascape platform. OpenScientist was provided with a CSV file containing this data along with enriched pathways and associated proteins and was asked to interpret the biological significance of the pathways in the brain and how their age correlations and model weight directions might inform mechanisms linking aging to amyloidosis. At the time of the OpenScientist analysis, these data and results were unpublished; they have since been released as a preprint.[15]

OpenScientist demonstrated good understanding of the dataset and accurately summarized several key statistics, including the proportion of proteins with positive versus negative model weights and the proportion of proteins increasing or decreasing with age. These matched values generated during manual analysis. The agent also produced a statistical summary of the dataset, including counts of enriched pathways and distributions of age-associated proteins. However, some reported statistics were not particularly informative for the biological question, such as mean age correlations or median model weights across proteins.

When summarizing enriched pathways, OpenScientist introduced several inaccuracies. Many Reactome pathways were renamed, making them difficult to match to the original dataset because pathway identifiers were not reported. Several q-values were also slightly incorrect. For example, the pathway Attenuation phase (R-HSA-3371568) had a q-value of −2.04 in the dataset but was reported as −1.9 by the agent. In some cases, pathways that would have ranked among the most enriched were omitted from the summary. OpenScientist also generated a table highlighting proteins appearing across multiple pathways. This analysis was largely correct, although one counting error occurred for RAD23B.

The agent arrived at several interpretations similar to those from manual analysis. Both analyses converged on a model in which aging is associated with coordinated upregulation of autophagy, membrane trafficking, and cellular stress response pathways that may represent compensatory responses that ultimately fail to prevent amyloid pathology. OpenScientist also identified 14-3-3 proteins in the CSF proteome as markers of neurodegenerative processes, although it did not elaborate on their diverse chaperone-like functions or potential roles in regulating tau aggregation and amyloid clearance as extensively as the human analysis.[16,17] The agent also proposed similar follow-up analyses, including testing whether these proteins predict future amyloidosis in longitudinal CSF samples and verifying whether CSF proteomic changes reflect underlying brain pathology, which was already underway by human researchers.

OpenScientist also generated several novel hypotheses that were not identified during manual review. The agent highlighted calpastatin (CAST) as a potential regulator of calpain activity and noted that calpain activation has been reported to precede tau phosphorylation, suggesting a possible mechanistic link between protease regulation and tau pathology.[18] It also observed that glycosyltransferases in the model all had positive weights, suggesting that increased glycosylation capacity might correlate with reduced amyloid pathology. This observation had been overlooked during manual analysis and represents a hypothesis for further investigation. The agent additionally suggested that changes in blood–brain barrier related proteins might reflect vascular aging processes influencing CSF proteomic shifts, although this interpretation was presented with limited supporting evidence.

OpenScientist completed the analysis in approximately one hour compared with several days required for manual interpretation. However, extensive verification was required because the system frequently hallucinated references. Of eight references listed, six contained incorrect titles or authors and none included DOIs, making them difficult to validate. Overall,

OpenScientist reproduced many of the same high-level conclusions reached through human analysis while identifying several additional hypotheses for follow-up. At the same time, the agent missed some mechanistic details and introduced data inconsistencies, highlighting the importance of expert review when using agentic AI systems for biological interpretation.

After completion of the formal evaluation period described in this manuscript, we performed an additional post hoc run of OpenScientist using an updated system configuration to determine whether several of the most significant issues observed in the initial run could be resolved. In this subsequent run, the agent correctly cited non-hallucinated literature with valid PMIDs linking to appropriate scientific articles, addressing the major reference reliability problem observed previously. The system also generated a well-structured summary table describing pathway-level trends, dominant model weight directions, age-correlation patterns, key proteins, and potential biological mechanisms, a format that in some respects was clearer than the tables used in the original human analysis. However, some issues remained. OpenScientist continued to rename Reactome pathways without reporting pathway identifiers or original enrichment scores, making it difficult to match entries directly to the input dataset. Despite these limitations, the updated run produced interpretations consistent with the initial analysis, suggesting that the agent's core biological conclusions were stable across runs and not strongly driven by stochastic variation.
